# Risk Factors for Mild, Moderate, and Severe Alcohol Use Disorder (AUD) in a sample of adult substance users: Implications for DSM-5 AUD Classification

**DOI:** 10.1101/2020.10.26.20219824

**Authors:** Zachary L. Mannes, Dvora Shmulewitz, Ofir Livne, Malka Stohl, Deborah Hasin

**Affiliations:** Department of Epidemiology, Columbia University Mailman School of Public Health, New York, NY; New York State Psychiatric Institute, New York, NY; Department of Psychiatry, Columbia University Medical Center, New York, NY

**Keywords:** Alcohol Use Disorder, AUD, Alcohol Use, Mental Health, Functional Impairment

## Abstract

Though risk factors of Alcohol Use Disorder (AUD) have been well-studied, information is lacking on whether clinical characteristics differentiate between the three levels of severity (mild, moderate, severe) that were established for the first time in the Diagnostic and Statistical Manual of Mental Disorders, fifth edition (DSM-5). Therefore, in this study, we examined the association between alcohol consumption, mental and physical health, and functional impairment with the three DSM-5 AUD severity levels among adults age 18+ (N=588) pre-screened for problems with at least one substance. Participants recruited between 2016-2019 completed measures of AUD, harmful alcohol use, psychiatric conditions, and mental, physical, and social functional impairment. For each predictor, a multinomial logistic regression model was used to evaluate the association with a four-level AUD outcome (mild, moderate, severe, vs none), controlling for sociodemographic characteristics and other substance use. Twelve-month prevalence of none, mild, moderate, and severe DSM-5 AUD was 34.0%, 12.2%, 13.4%, and 40.3%, respectively. Participants reported a mean of 11.3 (SD=9.90) days of alcohol use in the past month, nearly half (48.0%) perceived to have a major problem with alcohol, and 61.4% met the threshold for harmful drinking. Multinomial logistic regression demonstrated that compared to the reference group (no AUD), all three AUD severity levels were associated with drinking frequency, problematic, and harmful alcohol use. However, only severe AUD was associated with personality disorders: (AOR=1.91, 95% CI=1.28, 2.86), MDD (AOR= 2.44, 95% CI= 1.62, 3.66) or PTSD (AOR= 1.65, 95% CI= 1.00, 2.71),. Similarly, only severe AUD was associated with impaired physical (AOR= 1.63, 95% CI= 1.01, 2.61), mental (AOR= 1.80, 95% CI= 1.16, 2.79), and social functioning (AOR= 1.87, 95% CI= 1.39, 2.51). This study adds to existing literature on clinical correlates of AUD by further elucidating the risk factors of the different AUD severity groups, while also highlighting an important, differential observation wherein measures of psychiatric disorders and functional impairment were only associated with severe AUD. The study suggests that the DSM-5 category of severe AUD most closely corresponds to AUD cases often found in secondary or tertiary treatment settings, and that cases of mild or moderate AUD may warrant less intensive treatment approaches. Future investigations should seek to examine the validity of the DSM-5 AUD three-level severity distinction by using longitudinal designs to evaluate change in mental health and functioning over time, along with their association with AUD severity classification.

## Introduction

Alcohol consumption is prevalent in the United States with 66% of adults reporting use in the past year and 6% report heavy or high-risk drinking (Boersma, Villarroel, & Vahratian, 2018; Grant et al., 2017; Hasin et al., 2017). Excess alcohol use remains among one of the leading causes of preventable morbidity and mortality in the United States, and continues to account for a large proportion of substance-related fatalities (Kendler et al., 2016; Mokdad et al., 2004). Heavy drinking is a risk factor for developing Alcohol Use Disorders (AUD), which is characterized by a problematic and persistent pattern of alcohol use that contributes to significant impairment and distress (American Psychiatric Association [APA], 2013; Grant et al., 2015). AUD is associated with a multitude of medical and psychiatric comorbidities (Castillo-Carniglia, Keyes, Hasin, & Cerdá, 2019; Xi et al., 2017). AUD also increases the risk of mortality as a result of injury, cancer, and cardiovascular disease among other causes (Roerecke, & Rehm, 2014).

The diagnostic criteria for AUD have undergone considerable revision since the advent of the first Diagnostic and Statistical Manual of Mental Disorders (DSM) in 1952 (APA, 1952). The third edition, DSM-III, was the first version to create the abuse/dependence classification using specific diagnostic criteria (APA, 1980). Published in 1987, the DSM-III-R (revised) further modified the AUD criteria of substance abuse and substance dependence based on emprical research and recommendations by the World Health Organization (APA, 1987; Edwards, Arif, & Hadgson, 1981; Kosten et al., 1987; Rounsaville, Kosten, Williams, & Spitzer, 1987). Though DSM-IV largely maintained the AUD differentiation between abuse and dependence, AUD criteria were modified in DSM-5 based on epidemiological and clinical research that supported a dimensional approach to diagnosis compared with a categorical classification (Dawson et al., 2010; Hagman,& Cohn, 2011; Hasin & Beseler, 2009; Hasin et al, 2013). As a result, DSM-5 replaced the distinction between abuse and dependence with a single disorder defined by the same 11 criteria that had formerly been used to define abuse and dependence, except that craving was added and legal problems removed. Importantly, DSM-5 proposed that AUD be classified as mild (2–3 criteria), moderate (4–5 criteria), and severe (6+ criteria). This modification was intended to identify individuals with varying AUD severity and differences in the extent of impairment and disability. However, the clinical usefulness of the 3-level severity categorization has been debated (Fazzino et al., 2014a; Fazzino 2014b), including discussion of the utility of a mild AUD category with just two symptoms (Borgess et al., 2011; Goldstein et al., 2015). Recent preliminary evidence from a clinical trial indicated that DSM-5 AUD severity categories have clinical relevance among treatment seeking adults (Kiluk et al., 2018). However, this study only included 68 participants reporting AUD symptoms over the past 30 days (not the 12 months required for DSM-5 diagnosis), which may limit generalizability.

To better understand the validity and potential utility of the DSM-5 AUD severity differentiations, further information is needed on whether clinical characteristics differentiate between the three-level AUD severity groups (mild, moderate, severe). In particular, validity of the 3-level severity measure of DSM-5 AUD is unclear, as few studies have delineated the extent of alcohol use, impairment, and functioning across the three AUD severity groups. Therefore, the purpose of this study was to examine the association between a set of external validators, including alcohol consumption, mental and physical health, and functional impairment, with the DSM-5 AUD severity classifications among a heterogeneous sample of 588 adults reporting binge drinking and/or illicit substance use. We hypothesized that all validators would be associated with each AUD severity category. Moreover, we anticipated that the relationship between the validators and AUD severity would demonstrate a dose response relationship, such that the magnitude of the association between the validators and AUD severity would be greater for each additionally severe category.

## Methods

### Participants and Procedures

Methods of the study have been described in detail elsewhere (Hasin et al., 2020). Briefly, the sample consisted of 588 adults age 18 years or older recruited between 2016-2019 at a suburban addiction treatment program (n=150) and at an urban medical center (n=438). All participants reported binge drinking [i.e., ≥5 drinks or ≥4 drinks for men and women respectively] and/or illicit substance use (i.e., cannabis, cocaine, opioid use) in the past 30 days or the 30 days prior to inpatient substance use admission. Moreover, all participants reported ≥1 DSM-5 substance use disorder (SUD) criteria in the pre-study screening. Adults recruited at the inpatient program were informed of the study via hospital staff or posted flyers and after expressing initial interest, met with an on-site, Master’s level research coordinator that screened for eligibility criteria. Participants recruited at the medical center were informed of the study through two types of advertisements: 1) newspapers ads that invited individuals to call a research coordinator who then briefly explained the study and prescreened for eligibility criteria and 2) Facebook ads that offered a brief explanation of the study and provided a link to an online prescreening survey. After obtaining written informed consent, participants answered questions pertaining to demographics, substance use, mental health, and functional impairment. Adults were excluded from the study if they were non-English speaking, had a hearing, visual, or cognitive impairment that prevented participation, planned to relocate from the area, or were currently psychotic, suicidal, or homicidal. All participants were compensated $50 for the initial, baseline interview and $35 for the re-test (not discussed in this study). This study was approved by the Institutional Review Boards of New York State Psychiatric Institute and South Oaks Hospital.

### Interviewers, training and supervision

All interviewers had at least a master’s degree in psychology or social work and an average of 4.5 years of clinical experience (range, 1-10 years). PRISM-5 training included a manual, 2-day workshop, practice interviewing, role-playing, and certification. To become certified, trainees recorded mock interviews that underwent structured review by PRISM trainers. Trainees became interviewers after 2 recordings were rated satisfactory or better. Supervision of the study interviewers was conducted by trained, highly experienced trainer/supervisors with clinical masters degrees (psychology or social work) whose mean years of clinical experience utilizing the PRISM interview in research settings was 7.6 years (range: 3-10), and whose mean years of supervisory experience with the PRISM-5 was 6 years (range: 2-8). After certification, PRISM-5 supervisors met weekly with interviewers. They also reviewed recordings of 10% of the interviews, scoring interviewer performance, which occasionally indicated typical issues in such interviewing that required supervision (e.g., reading probes as written) but generally indicated that PRISM-5 interviews were largely conducted in a standardized way according to the PRISM-5 training procedures. Further information about the PRISM-5 and procedures used in this study can be found elsewhere (Hasin et al., 2020).

### Measures

#### Outcome

##### Alcohol Use Disorder

The PRISM-5, a semi-structured, computer-assisted interview, assessed the 11 DSM-5 AUD criteria among those who used alcohol at least 6 times within the past 12-months. DSM-5 AUD criteria include: 1) withdrawal, 2) tolerance, 3) increasing quantity or frequency of use/longer episodes of use, 4) persistent desire or unsuccessful attempts to decrease/control use, 5) lots of time spent obtaining, using or recovering from effects of alcohol, 6) social, occupational, or recreational activities given up or reduced because of use, 7) continued drinking despite knowledge of use causing or exacerbating a medical condition, 8) recurrent difficulties to fulfill major role obligations, 9) recurrent use in hazardous situations, 10) craving/strong desire to consume alcohol, 11) continued use despite interpersonal challenges caused by alcohol consumption. The PRISM-5 produces DSM-5 AUD diagnoses via computer algorithms and classified participants based on disorder severity: no disorder (0-1 criteria), mild (2-3 criteria), moderate (4-5 criteria) and severe (≥6 criteria) disorder. PRISM-5 AUD diagnostic measures demonstrate good to excellent reliability and validity among adults who consume alcohol (Hasin et al., 2020), and have been used as validators of non-clinician interviews as part of a national survey (Hasin et al., 2015).

#### Predictors

##### Alcohol Use Frequency and Consequences

Two measures of alcohol use problems were defined using questions from the Addiction Severity Index (ASI; McLellan et al., 1992). The ASI, widely used in research and clinical settings, is often utilized in self-administered format, as it was in the present study (Butler et al., 2001; Rosen et al., 2000). One ASI measure used was a continuous variable indicating the number days the participant drank alcohol in the past 30 days. The second measure was a binary variable indicating whether participants felt that alcohol had been a major problem for them. The third measure was based on the Alcohol Use Disorders Identification Test (AUDIT), a questionnaire that assesses the frequency of eight indications of harmful drinking on a 5-point scale (0= “Never” to 4= “4 or more time weekly”) and two additional questions assessing past-year alcohol associated injuries and familial/peer concern related to alcohol use (Allen et al., 1997; Saunders et al., 1993). In line with the scoring guidelines, participant responses were summed, with a score of 8 or more indicating harmful alcohol use in the past year. Participants were dichotomized based on this designation. The AUDIT is one of the most widely used measures of alcohol-associated difficulties, and demonstrates strong psychometrics across clinical populations (Daeppen et al., 2000; de Meneses-Gaya et al., 2009; Higgins-Biddle & Babor, 2018).

##### Social Functioning

The Social Adjustment Scale Self-Report (SAS-SR) is a widely used, reliable, self-report measure that assesses role functioning in the prior two weeks across areas of work/school, social/leisure time, family outside the home, primary relationship, parental role, and family unit (Weissman et al., 2001; Weissman & Bothwell, 1976; Weissman et al., 1978). Each item is assessed on a 5-point scale, with higher scores indicating greater impairment in functioning. In line with scoring guidelines, the scale was scored as the mean of the responses and the role areas not relevant to the respondent were skipped (Weissman & Bothwell, 1976).

##### Physical and Mental Health Functioning

The Medical Outcomes Study Short Form 12-Item (SF-12) measures functioning in eight domains (i.e., general health, physical functioning, role physical, bodily pain, vitality, social functioning, role emotional, and mental health; Ware, Kosinski, & Keller, 1996). Coinciding with the SF-12 scoring guidelines, a composite Physical and Mental Component Summary score was calculated from the subdomains. These summary scores indicated the extent to which physical, work, and social activities or accomplishments were negatively impacted by physical or mental health issues in the prior 4 weeks. These component scores are valid, reliable, widely used, and have been shown to be associated with DSM-5 AUD in several general population studies (Kirouac et al., 2017; Grant et al., 2015; Rubio et al., 2013). To facilitate interpretation for each component (physical, mental), impairment was defined as scoring in the bottom 25^th^ percentile, similar to other studies examining alcohol use (Aharonovich et al., 2017)

##### Mental Health Diagnosis

Four current disorders were included in the present study, Major Depressive Disorder (MDD), Post-traumatic stress disorder (PTSD), Borderline Personality Disorder (BPD), and Antisocial Personality Disorder (ASPD). The PRISM-5 was used for these assessments, with a full module for each disorder. Participants were categorized as currently having one of these disorders based on DSM-5 criteria for each diagnosis, including significant distress or functional impairment. For MDD, participants must have reported five or more symptoms during a 2-week period and at least one of the symptoms to be either (1) depressed mood or (2) loss of interest or pleasure. In order to meet a diagnosis for PTSD, participants had to endorse at least one symptom each from criterion A (i.e., direct exposure, witnessing event, learning of event, indirect exposure via traumatic details), criterion B (i.e., upsetting memories, nightmares, flashbacks, emotional distress, physical reactivity), and criterion C (avoidance of external reminders, thoughts, or feelings) and two symptoms from criterion D (anhedonia, isolation, negative affect, self-blame, difficulty recalling memories related to trauma) for at least one month. BPD required five of the nine criteria to be met across the domains of emotional dysregulation, impulsive behaviors, distorted perceptions of self, and unstable relationships over lifetime, with at least one symptom occurring in the past 12 months. ASPD required three or more criteria and onset of conduct disorder before the age of 15, lifetime, with at least one symptom occurring in the past 12 months. Two combined measures were also created, one indicating either of the personality disorders (BPD and ASPD), and one indicating any of the psychiatric disorders (BPD, ASPD, PTSD, or MDD). The PRISM-5 is commonly used to assess psychiatric conditions and demonstrates strong reliability and validity among adults reporting substance use (Hasin et al., 1996; Hasin et al., 2006; Hasin et al., 2015; Morgello et al., 2006; Ramos-Quiroga et al., 2012; Torrens et al., 2004).

##### Depressive Symptoms

The Patient Health Questionnaire (PHQ-9) was used as an alternative measure of depression. The PHQ-9 assesses self-reported depressive symptoms over the past two weeks, is widely used and has excellent reliability and validity, including in SUD populations (Kroenke, Spitzer, & Williams, 2001; Lowe et al., 2004; Delgadillo et al., 2011). Each of the nine items ranges from 0 “not at all” to 3 “nearly every day” with higher scores indicating more severe symptoms. Responses were summed, and the summary score was converted to a count variable as follows: 0-4 points (minimal or no problems) 5-9 points (mild problems), 10-14 points (moderate problems), 15-19 points (moderately severe problems), and 20+ points, (severe problems; Kroenke, Spitzer, & Williams, 2001).

#### Control variables

##### Sociodemographics

Control variables assessed in the PRISM-5 included: age (18-29, 30-39, 40-49, 50+), sex (male; female); race/ethnicity (non-Hispanic White; Hispanic, non-Hispanic Black, Asian, Native Hawaiian/Pacific Islander, American Indian or Native Alaskan); education (no college; at least some college); and participant type (inpatient sample; community sample).

##### Other substance use

For each other substance assessed in the PRISM-5 (tobacco, cannabis, cocaine, heroin, hallucinogens, other drugs, and non-prescription use of prescription opioids, stimulants, and sedatives), a variable was defined indicating past month use.

### Statistical analysis

Prevalences of the sociodemographic variables were calculated, overall and by AUD status. Prevalence was calculated for AUD and the categorical validators and means for the count validators. For each validator (predictor), a multinomial logistic regression model was used to evaluate the association of the validators with the four level outcome of AUD severity (mild, moderate, severe, vs none) while adjusting for age, sex, race/ethnicity, education, and participant type (model 1). Results are reported as adjusted odds ratios (aOR) for each severity level as compared to the reference of no disorder. To determine that association was not due to poly-substance use, regression models were re-run, also adjusting for other substances (tobacco, cannabis, cocaine, opioids, heroin, stimulants, sedatives, hallucinogens, other drug) use (model 2). Analyses were carried out using SAS 9.4.

## Results

### Sociodemographic characteristics and validators

The majority of the sample was male (69.7%), Black (47.7%), 50 and older (42.1%), unemployed (74.1%), unmarried (80.1%), and had attained a high school education or less (54.7%). Regarding alcohol consumption, participants reported a mean of 11.3 (SD=9.90) days of alcohol use in the past month, nearly half (48.0%) felt they had a major problem with alcohol, and 61.4% met the AUDIT-C threshold for harmful drinking. Prevalence of any past 12-month AUD was 65.9%. Twelve-month prevalence of none, mild, moderate, and severe AUD was 34.0%, 12.2%, 13.4%, and 40.3%, respectively. Participants identifying as male, 50 and older, Black, unemployed, unmarried and having a high school diploma or less reported the highest rates of AUD. Regarding mental health, the most prevalent conditions were as follows: MDD (39.3%), BPD (37.9%), ASPD (26.0%), and PTSD (20.1%), while approximately 2/3 (61.9%) of the sample met diagnostic criteria for any other psychiatric disorder. Participants reported a mean of 2.3 (SD=0.69) on the measure of social impairment. Please see Table 1 and 2 for more information pertaining to study descriptors and the sociodemographic makeup of the sample.

**Table 1:** Demographic characteristics of the sample by DSM-5 Alcohol Use Disorder severity level (N=588)

|  | DSM-5 use disorder severity |
| --- | --- |

|  | **No Disorder**<br>**(N=200)** | **Mild**<br>**(N=72)** | **Moderate**<br>**(N=79)** | **Severe**<br>**(N=237)** |
| --- | --- | --- | --- | --- |
| **Sociodemographic variable** | **% (n)** | **% (n)** | **% (n)** | **% (n)** |
| Participant Type |  |  |  |  |
| Inpatient | 34.0 (68) | 9.7 (7) | 10.1 (8) | 28.3 (67) |
| Community | 66.0 (132) | 90.3 (65) | 89.9 (71) | 71.7 (170) |
| Gender |  |  |  |  |
| Male | 68.5 (137) | 73.6 (53) | 68.4 (54) | 70.0 (166) |
| Female | 31.5 (63) | 26.4 (19) | 31.6 (25) | 30.0 (71) |
| Age |  |  |  |  |
| 18-29 | 29.5 (59) | 27.8 (20) | 13.9 (11) | 16.5 (39) |
| 30-39 | 19.0 (38) | 15.3 (11) | 13.9 (11) | 16.0 (38) |
| 40-49 | 15.5 (31) | 15.3 (11) | 24.1 (19) | 21.9 (52) |
| 50 | 36.0 (72) | 41.7 (30) | 48.1 (38) | 45.6 (108) |
| Race/Ethnicity |  |  |  |  |
| White | 32.0 (64) | 19.4 (14) | 13.9 (11) | 30.8 (73) |
| Black | 39.5 (79) | 58.3 (42) | 63.3 (50) | 46.4 (110) |
| Hispanic | 22.0 (44) | 13.9 (10) | 20.3 (16) | 17.7 (42) |
| Other | 6.5 (13) | 8.3 (6) | 2.5 (2) | 5.1 (12) |
| Education level |  |  |  |  |
| High School or less | 55.0 (110) | 54.2 (39) | 62.0 (49) | 52.3 (124) |
| College or more | 45.0 (90) | 45.8 (33) | 38.0 (30) | 47.7 (113) |
| Marital status |  |  |  |  |
| Not Married | 81.0 (162) | 87.5 (63) | 78.5 (62) | 77.6 (184) |
| Living together/ Married | 19.0 (38) | 12.5 (9) | 21.5 (17) | 22.4 (53) |
| Employment |  |  |  |  |
| No job | 73.0 (146) | 62.5 (45) | 84.8 (67) | 75.1 (178) |
| Any job | 27.0 (54) | 37.5 (27) | 15.2 (12) | 24.9 (59) |
| Housing |  |  |  |  |
| Group home or homeless | 9.0 (18) | 5.6 (4) | 5.1 (4) | 9.7 (23) |
| Not in a group home or homeless | 91.0 (182) | 94.4 (68) | 94.9 (75) | 90.3 (214) |

**Table 2:** Distribution of Validators (N=588)

|  | **n** | **%** |
| --- | --- | --- |
| *Alternate alcohol problem measures* |  |  |
| Considered alcohol use to be a major problem | 282 | 48.0 |
| AUDIT | 361 | 61.4 |
| *Other DSM-5 psychiatric disorders* |  |  |
| Any other psychiatric disorder | 364 | 61.9 |
| Any personality disorder | 273 | 46.4 |
| Antisocial personality disorder | 153 | 26.0 |
| Borderline personality disorder | 223 | 37.9 |
| Major depressive disorder | 231 | 39.3 |
| Post-traumatic stress disorder | 118 | 20.1 |
| *Functional impairment* |  |  |
| SF-12 physical functional impairment | 150 | 25.5 |
| SF-12 mental functional impairment | 147 | 25.0 |
| **Count validator** | **Mean (SD)** | **Range** |
| Number of days used alcohol in last 30 days | 11.3 (9.90) | 0 - 30 |
| Functioning: Social adjustment scale | 2.3 (0.69) | 1.0 - 4.6 |
| Dimensional psychopathology scale (PHQ-9) | 2.2 (1.20) | 1.0 - 5.0 |

### Characteristics Associated with Mild, Moderate, and Severe AUD

After controlling for gender, age, race/ethnicity, education level, and participant type (i.e., inpatient substance use treatment and outpatient medical center; Model 1) frequency of alcohol use was associated with AUD such that participants reporting more days of use during the past month demonstrated greater odds of mild (AOR= 1.11 95% CI= 1.06, 1.15), moderate (AOR= 1.18, 95% CI= 1.14, 1.23), or severe (AOR= 1.18, 95% CI= 1.14, 1.21) AUD than no disorder. Similarly, adults reporting problematic alcohol consumption evidenced greater odds of mild (AOR= 3.25, 95% CI= 1.74, 6.07), moderate (AOR= 7.89, 95% CI= 4.29, 14.53) or severe AUD (AOR= 11.87, 95% CI= 7.37, 19.12) compared to participants not reporting problematic alcohol use. In regard to mental health, a different trend was observed such that PRISM-5 diagnosis and depressive symptoms were not associated with mild or moderate AUD. However, participants meeting PRISM-5 criteria for MDD (AOR= 2.44, 95% CI= 1.62, 3.66) or PTSD (AOR= 1.65, 95% CI= 1.00, 2.71), and personality disorder(s) (AOR=1.91, 95% CI=1.28, 2.86), had significantly greater odds of having severe AUD compared to adults not meeting criteria for those conditions. Similarly, perceived social functioning and functional impairment (mental and physical) were not associated with mild or moderate AUD, though poorer social adjustment (AOR= 1.87, 95% CI= 1.39, 2.51) was associated with severe AUD, and participants reporting physical (AOR= 1.63, 95% CI= 1.01, 2.61), and mental functional impairment (AOR= 1.80, 95% CI= 1.16, 2.79) were more likely to report severe AUD compared to those not endorsing functional impairment (Model 1). After controlling for other substance use, all results remained largely consistent though the effect of physical functional impairment on severe AUD status was attenuated and had a confidence interval with a lower bound that included 1.00 (AOR= 1.64, 95% CI= 1.00, 2.68). Please refer to Table 3 for additional information related to the risk factors of mild, moderate, and severe AUD.

**Table 3:** Association of alcohol, psychiatric and functioning validators with DSM-5 Alcohol Use Disorder, by severity level

| Validator | Model 1<sup>a</sup> |  |  | Model 2<sup>b</sup> |  |  |
| --- | --- | --- | --- | --- | --- | --- |
|  | Mild | Moderate | Severe | Mild | Moderate | Severe |
|  | AOR (95% CI) | AOR (95% CI) | AOR (95% CI) | AOR (95% CI) | AOR (95% CI) | AOR (95% CI) |
| Alcohol validators |  |  |  |  |  |  |
| Number of days drank alcohol in prior month | 1.11 (1.06, 1.15) | 1.18 (1.14, 1.23) | 1.18 (1.14, 1.21) |  |  |  |
| Considered alcohol use a problem<sup>c</sup> | 3.25 (1.74, 6.07) | 7.89 (4.29, 14.53) | 11.87 (7.37, 19.12) |  |  |  |
| Alcohol Use Disorders Identification Test<sup>c</sup> | 4.92 (2.68, 9.05) | 16.68 (8.52, 32.65) | 76.70 (39.42, 149.24) |  |  |  |
| Psychiatric validators |  |  |  |  |  |  |
| Current (past 12 month) DSM-5 disorders<sup>c</sup> |  |  |  |  |  |  |
| Any other psychiatric disorder<sup>d</sup> | 1.23 (0.68, 2.20) | 1.23 (0.70, 2.16) | 2.67 (1.73, 4.13) | 1.21 (0.67, 2.21) | 1.24 (0.70, 2.21) | 2.77 (1.76, 4.35) |
| Any personality disorder<sup>e</sup> | 0.95<br>(0.53,<br>1.71) | 0.84<br>(0.47,<br>1.50) | **1.91 (1.28,**<br>**2.86)** | 0.94<br>(0.52,<br>1.71) | 0.84<br>(0.47,<br>1.52) | **1.89**<br>**(1.25,**<br>**2.87)** |
| Antisocial personality disorder (ASPD) | 0.59<br>(0.28,<br>1.28) | 1.11<br>(0.57,<br>2.13) | **1.78 (1.14,**<br>**2.77)** | 0.57<br>(0.26,<br>1.25) | 1.09<br>(0.55,<br>2.14) | **1.79**<br>**(1.13,**<br>**2.84)** |
| Borderline personality disorder (BPD) | 1.17<br>(0.64,<br>2.15) | 0.62<br>(0.32,<br>1.19) | **1.99 (1.32,**<br>**3.00)** | 1.14<br>(0.62,<br>2.11) | 0.62<br>(0.32,<br>1.20) | **1.97**<br>**(1.30,**<br>**3.01)** |
| Major depressive disorder (MDD) | 0.96<br>(0.51,<br>1.78) | 0.79<br>(0.42,<br>1.46) | **2.44 (1.62,**<br>**3.66)** | 0.89<br>(0.47,<br>1.68) | 0.77<br>(0.41,<br>1.45) | **2.31**<br>**(1.52,**<br>**3.53)** |
| Post-traumatic stress disorder (PTSD) | 1.44<br>(0.70,<br>2.97) | 1.91<br>(0.97,<br>3.77) | **1.65 (1.00,**<br>**2.71)** | 1.47<br>(0.70,<br>3.07) | 1.94<br>(0.97,<br>3.89) | **1.74**<br>**(1.04,**<br>**2.91)** |
| *Dimensional psychopathology scale (PHQ-9)<sup>f</sup>* | 0.77<br>(0.59,<br>1.02) | 0.98<br>(0.77,<br>1.25) | **1.41 (1.20,**<br>**1.67)** | 0.77<br>(0.57,<br>1.03) | 1.02<br>(0.79,<br>1.32) | **1.49**<br>**(1.25,**<br>**1.78)** |
| **Functioning validators** |  |  |  |  |  |  |
| Social Adjustment Scale<sup>g</sup> | 0.73<br>(0.46,<br>1.15) | 0.97<br>(0.63,<br>1.47) | **1.87 (1.39,**<br>**2.51)** | 0.72<br>(0.45,<br>1.15) | 0.99<br>(0.64,<br>1.53) | **1.87**<br>**(1.38,**<br>**2.54)** |
| SF-12v2 physical functional impairment<sup>c,h</sup> | 1.20<br>(0.60,<br>2.40) | 1.82<br>(0.98,<br>3.37) | **1.63 (1.01,**<br>**2.61)** | 1.22<br>(0.60,<br>2.48) | 1.86<br>(0.99,<br>3.51) | 1.64<br>(1.00,<br>2.68) |
| SF-12v2 mental functional impairment<sup>c,i</sup> | 0.43<br>(0.19,<br>0.97) | 0.43<br>(0.19,<br>0.99) | **1.80 (1.16,**<br>**2.79)** | 0.42<br>(0.18,<br>0.98) | 0.45<br>(0.20,<br>1.04) | **1.90**<br>**(1.20,**<br>**3.01)** |

<sup>a</sup> Estimated from logistic regression analysis, adjusted for gender, age, race/ethnicity, education level and participant type
<sup>b</sup> also adjusted for other substance use
<sup>c</sup> reference level is No
<sup>d</sup> BPD, ASPD, MDD, or PTSD
<sup>e</sup> BPD or ASPD
<sup>f</sup> PHQ-9 = patient health questionnaire-9, assesses low affect over the past two weeks, with 5 responses, no problems - severe problems;
<sup>g</sup> continuous measure from 1-5, higher values indicate worse functioning
<sup>h</sup> impairment is indicated by scoring in the lowest quartile of the SF-12 physical component summary score
<sup>i</sup> impairment is indicated by scoring in the lowest quartile of the SF-12 mental component
summary score

## Discussion

This study examined DSM-5 AUD symptoms and their association with AUD severity classification (i.e., mild, moderate, severe) in a large, representative cohort of 588 adults reporting past 30-day alcohol and illicit substance use. Our hypotheses were partially supported, as the indicators of alcohol use severity such as number of days using alcohol in the past month, perception of problematic alcohol use, and harmful use were associated with all classifications of AUD severity. Moreover, the magnitude of the effect of the three alcohol measures strengthened with increasing AUD severity. However, a different pattern emerged for co-occurring psychiatric conditions and impaired role functioning in physical, mental, and social aspects, which were only associated with the severe level of AUD. This study builds on the literature by further elucidating the risk factors of mild, moderate, and severe AUD, while also highlighting an important, differential observation wherein no measures of functioning were associated with mild or moderate AUD.

The rates of DSM-5 AUD in this study were similar to those observed in another recent study (Kiluk et al., 2018). In our sample, adults diagnosed with severe AUD experienced pronounced impairment in the areas of mental health and psychosocial functioning relative to those with none, mild or moderate AUD. This finding is in line with literature demonstrating that increasing AUD severity is associated with worse social and role functioning, as well as psychiatric comorbidities. Specifically, adults with severe AUD have a greater likelihood of meeting diagnostic criteria for MDD, ASPD, and BPD compared to individuals with a mild or moderate disorder (Grant et al., 2015). Moreover, adults meeting PTSD criteria in this sample also demonstrated greater odds of severe AUD, contrasting previous findings (Grant et al., 2015). The nature of the relationship between psychiatric conditions and AUD is complex. AUD may lead to the development of psychopathology via escalating use and subsequent functional and psychosocial impairment (precipitation hypothesis). Conversely, psychiatric conditions may lead to AUD via consuming alcohol to remediate mental health symptoms (self-medication hypothesis), In addition, shared genetic and environmental causes contribute to vulnerability of both AUD and psychiatric conditions (Castillo-Carniglia, Keyes, Hasin, & Cerdá, 2019). The cross-sectional nature of this study precludes conclusions regarding the temporality of the observed associations, but regardless of the direction of effect, the associations of psychiatric problems and impaired functioning with AUD were only observed among participants whose AUD was at the severe level.

All three AUD severity classifications were associated with alcohol frequency and perceived problems with alcohol, though harmful alcohol use (AUDIT ≥8) demonstrated the strongest effect with severe AUD. These findings suggest that frequency and quantity of alcohol use may be insufficient for differentiating between the various AUD severity levels, while functionality may be the most salient indicator of AUD severity. This is intuitive given that people with severe AUD should endorse greater difficulty across a spectrum of functional symptoms. These findings are in line with Kiluk et al., 2018 investigation of alcohol drinkers showing that current alcohol use (quantity and frequency) was not associated with AUD severity, further highlighting that alcohol use alone is not adequate for diagnosing AUD, nor a sufficient measurement of AUD improvement in treatment. Additionally, those not meeting the diagnostic threshold for AUD (<2 symptoms) at the end-of-treatment were not significantly different from those with mild AUD severity in terms of alcohol abstinence, heavy drinking, or negative consequences, supporting previous research showing that the threshold of two criteria for AUD diagnosis is too low and may not represent “true cases” of AUD (Kiluk et al., 2018; Goldstein et al., 2015).

The debate pertaining to the minimum clinical cutoff of DSM-5 AUD and its operationalization is ongoing and has led to attempts to modify and optimize AUD diagnostic criteria (Fazino et al., 2014a). To this end, there is no current universal consensus for a new diagnostic assessment of AUD. Research has supported utilizing a continuous, criteria count measurement of AUD severity while others have proposed alternative severity cut points (Borges et al., 2011; Fazino et al., 2014b). Findings from this study lend support to the validity of DSM-5 severe AUD, while further research is needed to examine the extent of impairment among adults experiencing mild or moderate AUD.

Study limitations are noted. First, our study utilized self-report measures of substance use and mental health symptoms (PHQ-9) and participants may have under-reported their symptoms. Moreover, reporting on indications of “problematic” alcohol use may necessitate awareness, recognition, and vulnerability that are lacking among participants not yet willing to acknowledge their issues with alcohol use. Secondly, the study utilized a cross-sectional design; thus, we cannot determine the temporality of the observed relationships. Thirdly, approximately one-fourth of the participants were currently in treatment for a SUD, and thus were likely to currently abstain from using alcohol. These individuals were also likely receiving mental health services, which may have improved symptomatology and subsequent scores on the PHQ-9, a measure of past two weeks depressive symptoms. However, the examined validators in this study were associated with the corresponding AUD severity groups even after controlling for recruitment location (i.e., inpatient treatment vs a major medical center) and subsequent treatment utilization. Despite these limitations, this study highlights important correlates of varying AUD severity levels, and elucidates potential diagnostic challenges with mild and moderate AUD. This study benefited from use of PRISM-5 diagnoses for AUD and psychiatric conditions, and also from use of a large cohort of binge alcohol users.

In conclusion, our study demonstrated that only severe AUD was associated with functional impairment and current psychiatric comorbidity. Moreover, problematic and harmful alcohol use patterns were most strongly associated with severe AUD. These are characteristics often found among patients in treatment for AUD in tertiary care, suggesting that severity levels of AUD might eventually be used to suggest intensity of interventions for AUD, e.g., brief interventions for those at mild or even moderate levels, with referral to more intensive treatment programs for those with severe AUD, as outlined in the SBIRT model (Substance Abuse and Mental Health Services Administration, 2020). This study builds on the existing literature by identifying salient risk factors of AUD, while also elucidating clinical differences in mental health and functional impairment such that all measures of functioning were only associated with severe AUD. Findings from this study also highlight the importance of examining AUD as a three-level construct instead of a dichotomous measure indicating presence of any AUD vs. none, an ongoing limitation in empirical studies (Takahashi et al., 2017; Wakefield & Schmitz, 2014). Given findings from this study, future investigations should seek to examine the criterion validity of the DSM-5 AUD three-level severity distinction by using longitudinal designs to evaluate change in mental health and functioning over time, along with their association with AUD severity classification.

## Data Availability

Data for this study was collected as a part of a larger study investigating DSM-5 Substance Use Disorders (R01DA018652). For questions regarding data, please contact

